# Combinatorial benefit without synergy in recent clinical trials of immune checkpoint inhibitors

**DOI:** 10.1101/2020.01.31.20019604

**Authors:** Adam C. Palmer, Benjamin Izar, Haeun Hwangbo, Peter K. Sorger

## Abstract

Hundreds of clinical trials are testing combinations of Immune Checkpoint Inhibitors (ICIs) with other cancer therapies in the hope that they will have additive or synergistic efficacy involving mechanisms such as immune priming. However we find that the clinically observed benefits of recently reported and approved combination therapies with ICIs are fully and accurately accounted for by increasing the chance of a single-agent response in individual patients (drug independence), with no requirement for additive or synergistic efficacy (correlation between observed and expected Progression Free Survival: Pearson r = 0.98, *P* = 5×10^−9^, n = 4173 patients in 14 trials). Thus, the likely anti-tumor efficacy of new ICI combinations can be predicted if monotherapy data are available; predicting adverse effects remain challenging. Realizing the promise of drug additivity or synergy is likely to require better response biomarkers that identify patients in whom multiple constituents of a combination therapy are active.

## INTRODUCTION

The introduction of immunotherapy is one of the most important recent developments in oncology. Immune checkpoint inhibitors (ICIs), particularly those directed against PD-1, PD-L1 and CTLA4, have improved survival for many but not all types of cancer^1^. To increase rates and durability of responses to these ICIs, hundreds of combination clinical trials are currently underway involving a wide range of drug classes^2^. A key hypothesis motivating these trials is that targeted drugs, cytotoxic chemotherapies and immunotherapies themselves can enhance responsiveness to ICIs by increasing tumor immunogenicity^3^ or by other less well understood mechanisms^4^. An anticipated outcome is that individual patients will experience a superior response to a combination than to either drug alone as a consequence of pharmacological additivity or synergy. However, as Emil Frei described in 1961^5^, drug combinations can also confer clinically meaningful benefit through independent action^6,7^, in which patient *populations* benefit (as measured by population-averaged outcomes, such as response rate or median Progression-Free Survival - PFS) even without additive or synergistic benefit to *individual* patients.

In a drug combination that improves on monotherapy via independent action, individual patients benefit primarily or entirely from whichever single drug is most active for them^6,7^. A combination provides a statistically significant benefit at the population level because each patient has more than one opportunity for a meaningful response to a single drug in the combination. This contrasts with additive or synergistic interaction in which drugs work together at the level of individual patients. Previous work has shown that independent action is broadly relevant to anti-cancer therapy in part because, in practically all types of cancer, patients vary widely in their sensitivities to individual drugs, and because responses are weakly correlated within and across drug classes^6^. Improvements conferred by combination therapies exhibiting independent action can be understood as a form of ‘bet-hedging’. Using current disease classifications and biomarker-based stratification, predicting which drug is best for an individual patient remains uncertain. A combination hedges this uncertainty by administering multiple drugs and thereby increases the chance of a single strong anti-tumor response. This concept is less familiar today than drug synergy but the earliest human trials of combination chemotherapy calculated the expected benefit from independent action and found that it accurately accounted for improvements in response rates^5,8,9^.

Clinical evidence for additivity or synergy is provided when the activity of a combination therapy surpasses the null hypothesis of independent action, as calculated from single-agent data (independent action is also an appropriate null hypothesis based on the Akaike Information Criterion, AIC^6^). We have previously analyzed responses to single and combined cancer therapies in multiple Phase III clinical trials, and in hundreds of patient-derived tumor xenografts, and found that many approved combination therapies provide a magnitude of survival benefit (measured by PFS) that is fully accounted for by drug independence^6^. A minority of approved combinations exhibited additivity or synergy by this definition, particularly when bevacizumab is combined with chemotherapy for metastatic cancer. It is not possible to distinguish additivity from synergy using the methods described here and we therefore test whether a trial exceeds the expectations of drug independence, based on an inference of best ‘single-agent’ activity from trials of the constituent drugs; if it does, the null hypothesis is rejected in favor of drug interaction (i.e. additivity or synergy). In the case of combinations of three or more drugs, ‘single-agent’ can be shorthand for a constituent treatment that is itself a combination of multiple drugs (e.g. etoposide plus platinum as the ‘chemotherapy’ arm of the IMpower133 trial in Small-Cell Lung Cancer^10^).

Our previous analysis of independent action chiefly involved combinations of targeted therapies and chemotherapies^6^ as phase III data were available for only one ICI combination: Ipilimumab (a CTLA-4 mAb) plus Nivolumab (a PD-1 mAb) in metastatic melanoma. We found that the duration of response (as measured by PFS) as well as changes in tumor volume were precisely consistent with independent action. Additional evidence for independent action in PD-1 inhibitor combinations was recently reported by Schmidt *et al*^11^ based on analysis of objective response rates. In this paper we analyze PFS data from all recent clinical trials of ICI combination therapies for which data on single agent and combination arms are available and look for evidence of additive or synergistic drug interaction.

## RESULTS

We analyzed thirteen recently reported Phase III trials of combinations involving ICIs in melanoma, squamous and nonsquamous non-small-cell lung cancers (NSCLC), small-cell lung cancer, renal cell cancer, triple-negative breast cancer, gastric and gastroesophageal junction cancer, and head and neck squamous cell carcinoma as well as a Phase II trial in BRAF-mutant melanoma^10,12–24^ (Supplementary Tables 1-3). Eleven of these trials led to approval by the Food and Drug Administration (FDA) of practice-changing therapies. As described previously^6^, we digitized Kaplan-Meier curves of PFS data for combination therapies and their constituents, and computed an expected curve under the null hypothesis of independence by randomly sampling single-agent PFS durations (with correlation ρ=0.3±0.2; Methods), and assigning each simulated patient the longer of the two sampled PFS values (**Figure 1**). Two trials in small-cell lung cancer of etoposide plus platinum with or without PD-L1 inhibition were merged for analysis^10,19^. We also updated our analysis of ipilimumab plus nivolumab^6^ in melanoma, for which longer follow-up data (42 months) are now available^12^.

**Figure 1.**
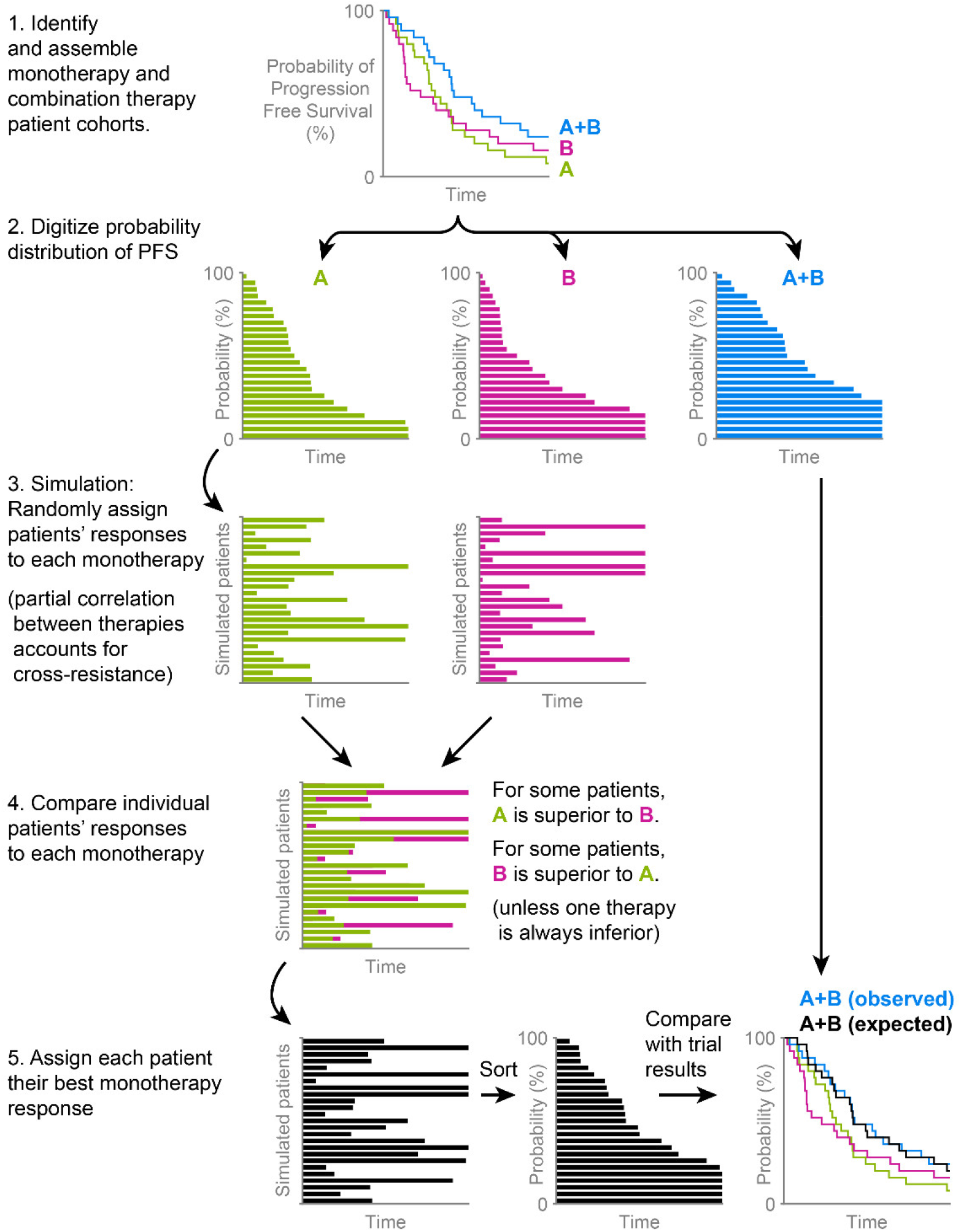
Calculating the expected activity of a combination therapy under the null hypothesis of independent drug action. Solely for clarity of illustration, quanta of probability are drawn as large (4%), and simulated patients are few (25); in practice PFS is resolved to <0.1%, and >10^4^ patients are simulated. Step 4 illustrates how patient-to-patient variability in best single-agent responses causes combination therapy to improve average response, but this does not occur if one therapy is so inferior to the other that it is never a better option for any individual patient.

We found that eleven trials exhibited PFS distributions very close to the prediction of the independence model (**Figure 2a-j**). In the case of the IMpower150 trial in nonsquamous NSCLC^24^, observed PFS exceeded the prediction of independence and in the KEYNOTE-022 trial in BRAF-mutant melanoma^21^ it was worse, as discussed in detail below. For the KEYNOTE-048 trial in head and neck squamous cell carcinoma^22^ and KEYNOTE-062 trial in gastric and gastroesophageal junction cancers^18^, published data made it possible to analyze cohorts with or without enrichment for PD-L1 expression. We found that PFS data for both PD-L1-enriched and non-enriched populations conformed to the expectations of drug independence (**Figure 2i, j**). Thus, even though biomarker-based stratification increases average response, it does not select for patients in which drugs interact pharmacologically.

**Figure 2.**
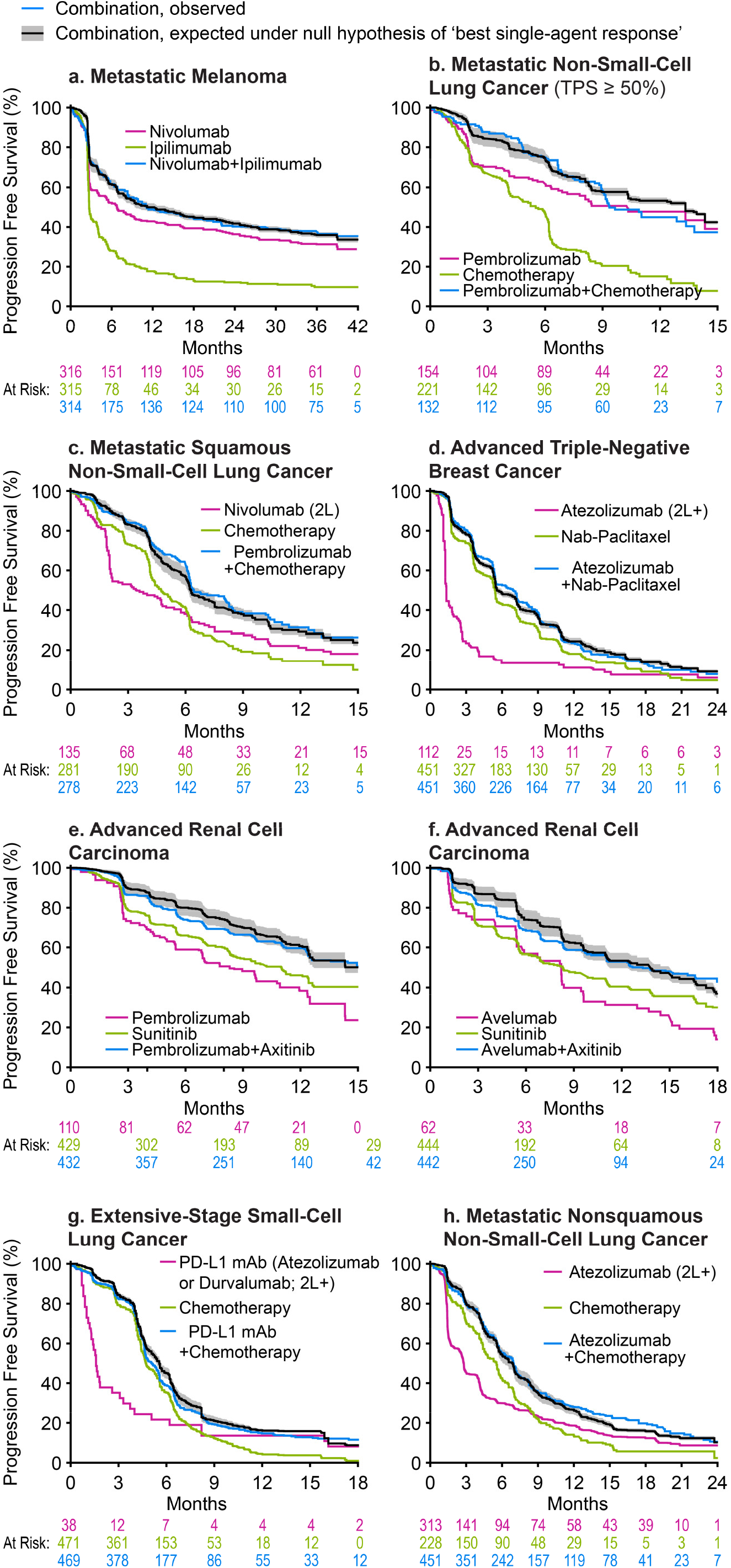
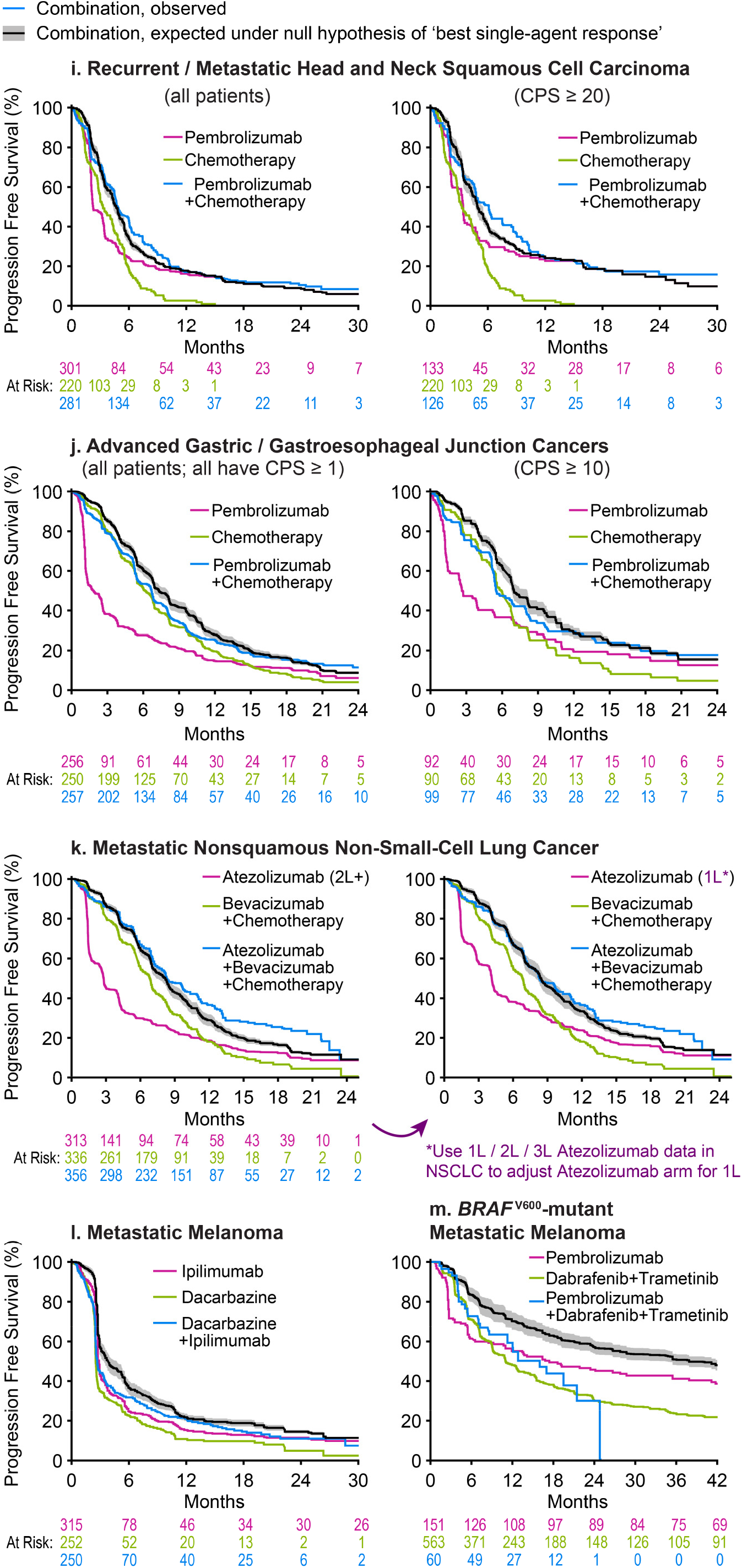
Progression Free Survival for combination therapies as observed in clinical trials and as predicted from independent activity of the therapies comprising the combination. Progression Free Survival (PFS) observed for each combination therapy (blue) was compared to that expected from the PFS distributions of the constituents of the combination (green and magenta) under the null hypothesis that each patient’s PFS is the best of their two possible responses to constituent therapies (black line and grey range, which reflects uncertainty in response correlation (ρ = 0.3 ± 0.2) or cross-resistance). 2L and 2L+ indicate data from patients treated at second-line or later; all other data are from patients previously untreated for metastatic or advanced cancer. TPS: PD-L1 Tumor Proportion Score. CPS: PD-L1 Combined Proportion Score. Combination therapy data from: **a** CheckMate 067^12^, **b** KEYNOTE-189^13^, **c** KEYNOTE-407^14^, **d** IMpassion130^17^, **e** KEYNOTE-426^16^, **f** JAVELIN Renal 101^15^, **g** IMpower133^10^ and CASPIAN^19^, **h** IMpower130^23^, **i** KEYNOTE-048^22^, **j** KEYNOTE-062^18^, **k** IMpower150^24^ (note that the difference between the prediction of independence – black line – and observed PFS – blue line - is significant for left panel and not for right panel; see Figure 3), **l** NCT00324155^20^. **m** KEYNOTE-022^21^. Data sources, patient characteristics, and limitations are described in Supplementary Tables 1 and 2.

To quantify similarity or difference from independence, we computed the hazard ratios attributable to additive or synergistic activity by digitally reconstructing individual patient events from Kaplan-Meier curves^25,26^. Patient data were then compared with PFS distributions predicted from independence. All combination therapy trials but IMpower150 were consistent with, or inferior to, the null hypothesis of independent action, as demonstrated by hazard ratios near 1.0 (**Figure 3a**). Moreover, observed PFS and expected PFS under the assumption of independence exhibited a very strong linear correlation, as illustrated in **Figure 3b** for a ‘landmark’ of 12 months on treatment (Pearson correlation r = 0.98, *P* < 10^−8^, n = 13 trials). When the same question was asked of PFS at all observed times, except for BRAF-mutant melanoma which was an inferior outlier, Pearson correlation was 0.99 and the mean absolute error of the independence model was less than 3%, which is within the margins of error of the trial data (**Figure 3c**; in this case, analysis of time-series data –which are not independent-preclude calculation of a *P* value). These findings constitute strong evidence that the dominant mode of benefit provided by ICI combinations involves independent drug action.

**Figure 3.**
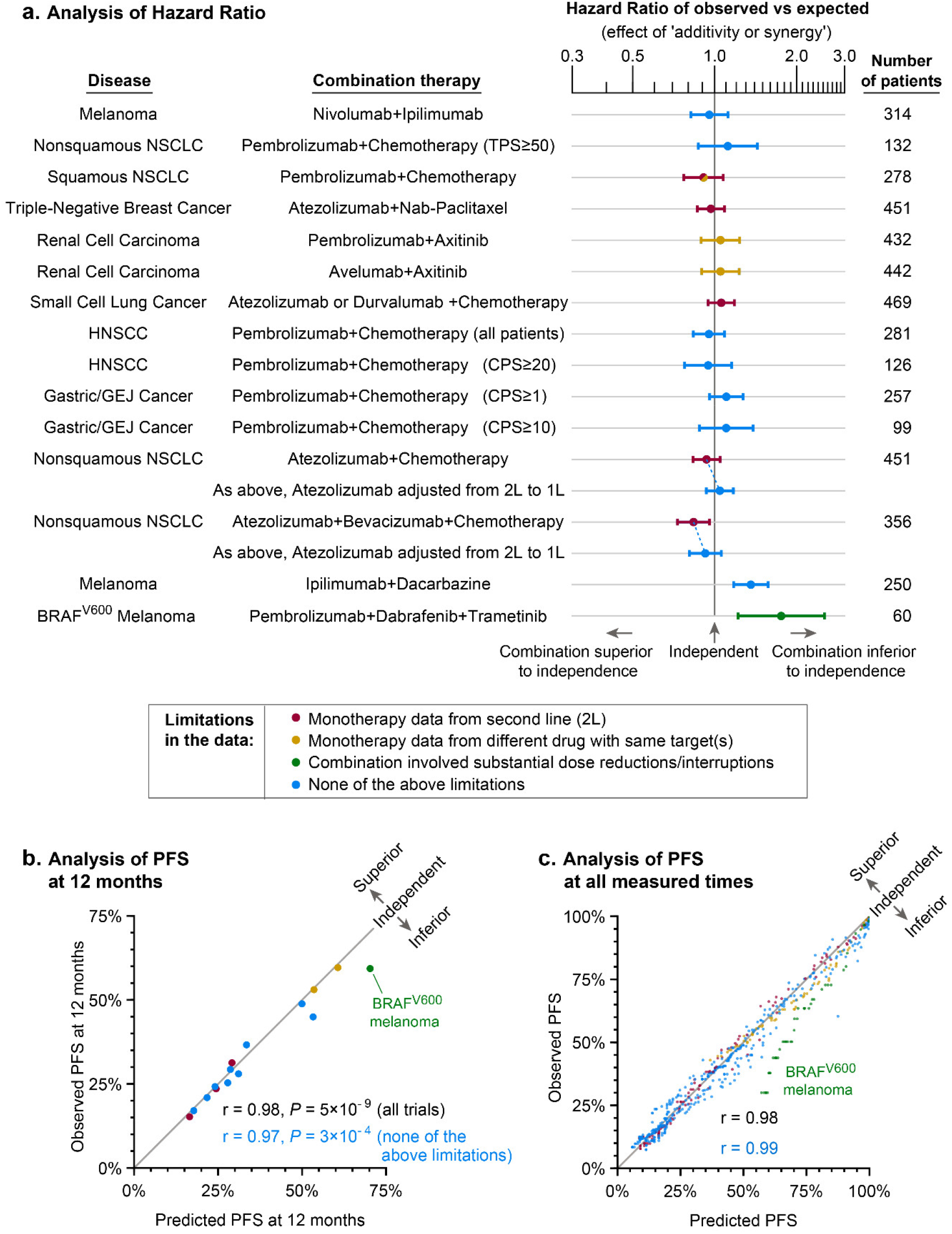
Trials combining Immune Checkpoint Inhibitors with other cancer therapies are consistent with, or inferior to, independent drug action. **a**. Patient events (progression or censoring) from clinical trials were compared to the prediction of independent drug action by computing the Hazard Ratio (Cox Proportional Hazards model). Error bars are 95% confidence intervals. Superiority to independence would indicate ‘drug additivity or synergy’, while inferiority could occur for multiple reasons including antagonistic interaction, toxicity necessitating dose reduction/interruption, or strong cross-resistance. HNSCC: Head and Neck Squamous Cell Carcinoma; NSCLC: Non-Small-Cell Lung Cancer; TPS: PD-L1 Tumor Proportion Score; CPS: PD-L1 Combined Proportion Score. Data in this and subsequent panels are color coded to denote deviation from the ideal case of a combination in which directly comparable monotherapies are available. See text for details. **b, c**. Observed PFS has a strong linear correlation with PFS expected under the null hypothesis of independent drug action, both at a landmark of 12 months (Pearson’s r = 0.98, *P* < 10^−8^, all trials) and over all measured times; r = 0.99 for trials excluding KEYNOTE-022^21^ of pembrolizumab + dabrafenib+trametinib for BRAF^V600^ melanoma, which was inferior to independence and for which dosing violated model assumptions (green points).

### Analyzing a possible case of additive or synergistic drug interaction in nonsquamous NSCLC

The IMpower150 trial evaluated first-line treatment of nonsquamous NSCLC by a combination of bevacizumab plus carboplatin plus paclitaxel with or without the PD-L1 inhibitor atezolizumab^24^. PFS in the ICI-containing arm exceeded the expectation of independence with a hazard ratio of 0.84 (*P* = 0.01, n = 356; the benefit attributable to drug interaction had a median PFS effect of +9 days; **Figures 2k, 3a**). Analysis of this trial was more complex than any other and we describe it in detail because it highlights challenges in the analysis of rapidly developing therapeutic areas in which single-agent data are not always available. IMpower150 did not evaluate atezolizumab alone and the only available data on atezolizumab monotherapy in nonsquamous NSCLC without PD-L1 pre-selection comes from the OAK trial^27^, which enrolled patients receiving their second or third line of treatment. However, a different trial of atezolizumab in NSCLC of all histologies (BIRCH trial; 72% nonsquamous, all tumors ≥5% PD-L1-positive)^28^ demonstrates that atezolizumab is more active as first-line than as second- or third-line therapy (PFS data was available in conference slides^29^). Thus our test of independence likely underestimated atezolizumab’s single-agent activity. To address this we used clinically observed differences in atezolizumab activity by line of therapy^28^ to construct a synthetic arm - as recently discussed for NSCLC^30^ - of first-line atezolizumab for nonsquamous NSCLC (**Figure 2k**; **Supplementary Figure 1**) and found that IMpower150 now closely matched the expectation of independence (Hazard Ratio 1.04, *P* = 0.46, n = 356; **Figure 3a**, dotted line denotes adjusted hazard ratio).

We are therefore left with two competing hypotheses about the four-drug combination tested in IMpower150: (a) atezolizumab is slightly less active at second-line than first-line, and independent action explains the benefit of adding atezolizumab to bevacizumab plus carboplatin plus paclitaxel, (b) atezolizumab is equally active at first and later lines of therapy and drug additivity or synergy is observed. Because atezolizumab is known to be more effective at the first-line of therapy in other NSCLC cohorts^28^ we conclude that hypothesis (a) is more likely. A more exact test of independence in cases such as this is likely to be impossible because combination therapy is so much better than monotherapy that the latter can no longer be ethically tested as a first-line option.

### Inferiority to independent action in a combination therapy for melanoma

The KEYNOTE-022 trial of BRAF plus MEK inhibitors (dabrafenib plus trametinib) with or without pembrolizumab in BRAF-mutant melanoma^21^ stood out as a case in which observed PFS was substantially worse than predicted by independence (Hazard Ratio=1.75, *P*=0.0025, n=60). The inferiority of the triple therapy may arise from the fact that nearly all patients required dose reduction, interruption, or discontinuation due to treatment-related adverse events. Dose reductions or interruptions can compromise the efficacy of each constituent of a regimen and negate the benefit of bet-hedging. This is likely of greatest concern when toxicity from an agent providing little benefit to an individual patient compels dose-reduction in an agent that is effective for that patient. This interpretation finds support in the analysis of volume change data from KEYNOTE-022, where initial responses in terms of volume change are approximately as good as independence predicts (**Supplementary Figure 3**; method from ^6^), so inferiority is observed only in durability of response. The benefit potentially achievable in BRAF-mutant melanoma using three agents (as predicted by independent action) could in principle be achieved by optimally stratifying patients to PD1 or BRAF plus MEK inhibition, making development of a predictive biomarker(s) for PD1 or kinase inhibitor response a priority in this disease.

### Limitations in the analysis

Retrospective analysis and trial reconstruction as performed here has inherent methodological limitations. First, the benefit conferred by a combination exhibiting independence varies with the correlation in response to individual agents; we have therefore used a range that accounts for partial cross-resistance^6^ based on data obtained on many drugs in PDX models (albeit not ICIs) and clinical experience that ICIs are not strongly cross-resistant with chemotherapies and other small molecule therapies. Given the limitations of mice in reproducing human immunotherapy responses, we note existing clinical evidence for low cross-resistance between ICIs and other therapies: among patients who have progressed on non-ICI first-line therapy, second-line ICI therapy achieves response rates that are only modestly lower than first-line ICI^28,31–33^. Strong cross-resistance would imply that after the failure of a first-line therapy, tumors would also be resistant to the second therapy, which is evidentially not the case for second-line ICIs. To account for uncertainty in the degree of cross-resistance, we compute independence for a range of correlation values ρ, which gives rise to a range of predicted PFS outcomes (**Figure 2**; gray range; mean width 4%, inter-quartile range 2% to 6%).

Second, the impact of differences in patient demographics on our analysis are largely unknown because demographically-linked response data are generally not published. Aggregate demographics are available and we checked that they were similar across matched monotherapy and combination therapy trial arms (**Supplementary Table 2**). However we cannot perform the sophisticated confounder studies being applied to real world data and to synthetic control arms^34,35^ with linked information on demographics and response; we encourage sponsors to perform this analysis themselves.

A third limitation is that many of the trials we studied did not include a monotherapy arm for the ICI under study, requiring the use of response data from another - typically earlier - trial testing the same ICI in a comparable patient cohort (**Supplementary Tables 1 and 2**). In these cases we matched the line of therapy and dosing to the greatest extent possible. Crucially, we permitted no substitutions or imperfections in the data that could bias against the discovery of additivity or synergy, but tolerated some biases in an opposite direction that could produce false discoveries of synergy. These cases are described using a color code in **Figure 3** and are described in detail below. In all cases we eliminated from consideration monotherapy data at higher doses or in healthier cohorts than combination therapies, because this could bias against the discovery of additivity or synergy.

In two cases (labelled orange in **Figure 3**) monotherapy data are not available for one constituent of a combination but instead for a similar drug known not to be clinically inferior. The first case is the KEYNOTE-407 trial in squamous NSCLC^14^ (**Figure 2c**), where single agent data is not available for anti-PD1 antibody pembrolizumab. The anti-PD1 antibody nivolumab is suggested as a non-inferior comparator by a meta-analysis of 1887 patients having NSCLC: no significant difference in PFS or overall survival between pembrolizumab and nivolumab was observed^36^. Thus, in the analysis of KEYNOTE-407, it is justifiable to use data on nivolumab in squamous NSCLC from CheckMate-017^37^ (note that CheckMate-017 studied second-line treatment which may bias in favor of synergy; see below). The second case occurs in renal cell carcinoma trials KEYNOTE-426^16^ and JAVELIN Renal 101^15^, in which the PDGFR/VEGFR/c-Kit receptor tyrosine kinase inhibitor axitinib was employed in the combination arm and the PDGFR/VEGFR/c-Kit receptor tyrosine kinase inhibitor sunitinib in the monotherapy arm. Fortunately, the assumption that axitinib is not inferior to sunitinib is supported by real-world clinical data^38^.

In five cases ICI monotherapy data were obtained from previously-treated (rather than untreated) patients (arms labelled ‘2L’ and data colored red; this includes IMpower150 described above). This biases us to overestimate - not underestimate - the likelihood of additivity or synergy, since cancer treatments are generally less, not more, active in second-line than first-line (because treatment promotes drug resistance). Thus, drug independence is almost always the more conservative model, not only from the perspective of AIC, but also when real limitations in available data are considered. If we consider only trials in which no compromises were necessary when comparing combination and monotherapy data the correlation between data and the model of independence remains r = 0.97 and *P* = 3×10^−4^ for PFS at 12 months. We conclude that our findings remain statistically significant under multiple scenarios of data inclusion and exclusion.

## DISCUSSION

Based on retrospective analysis of 13 combination immunotherapy Phase III trials in eight types of cancer we conclude that there is no clinical evidence of synergistic or additive interaction among ICIs or ICIs and other drugs. Instead, combinations examined to date are as beneficial as expected under the null hypothesis of independent drug action, including many combinations that have become first-line standards of care and are considered practice-changing. Thus, independent action can confer significant and highly meaningful clinical benefit through a process analogous to bet hedging. This conclusion is robust to the imperfections that are inevitably associated with retrospective analysis of clinical trial data, including the need to adjust for differences in first and second line-responses, the lack of data on some monotherapies, and the absence of detailed information on patient demographics. Sponsors in possession of the original clinical data are in a position to study the implications of independent action more thoroughly, with the resulting benefit that they will be better able to predict the likely outcome of future trials.

There is great interest in the potential of immunotherapies to confer highly durable benefits analogous to the curative chemotherapy combinations developed for several blood cancers^39^. Because single chemotherapies are almost never curative, cures in principle require combinations exhibiting additive or synergistic activity (for example, the RCHOP regimen used for lymphoma is additive)^40^. Used individually, ICIs have been remarkable in achieving long-lasting ‘treatment free survival’ as single agents^12,41^. As such, synergistic interaction is unnecessary for ICIs to elevate long-term survival as new ingredients in combinations with established therapies, provided that an ICI exhibits single-agent activity in the disease of interest. Is it nonetheless possible that longer follow-up of ICI trials might reveal drug additivity or synergy that is not yet evident? When follow-up has been sufficient for PFS to reach a low value (e.g. ≈20% of patients not progressing) and independent drug action is a sufficient explanation for benefit, we can conclude that additivity or synergy is absent in at least ≈80% of patients; this is true of seven trials examined. In three trials one-third or more of patients were not progressing at the longest times reported (15 to 42 months), raising the possibility of as-yet unobserved drug interaction. The possibility is strongest for pembrolizumab plus chemotherapy in PD-L1-high metastatic nonsquamous NSCLC^13^, and pembrolizumab or avelumab plus axitinib for advanced renal cell carcinoma^15,16^; longer follow-up will settle this issue. In the case of ipilimumab plus nivolumab for metastatic melanoma, 37% PFS was observed at three years, but tumor volume changes were consistent with independence across the entire patient population^6,42^ and sustained monotherapy responses continue to explain sustained combination therapy responses.

Widespread evidence of drug independence in ICI trials appears to conflict with pre-clinical data on immunotherapy combinations, including the concept of immune priming. Why might this be true? One possibility is that drug interactions at a molecular level^43^ do not prolong patient survival, which is the only response evaluated in this work. Drug interaction at a molecular level is not incompatible with drugs conferring independent survival benefits. Another possibility is that tumor volume changes measured in animal models do not correlate well with durability of response (PFS) in humans. However, we previously observed that both volume changes and PFS exhibit independence in most xenografts. Moreover, we note that in BRAF-mutant melanoma, near-term response (measured by volume change) is consistent with independence but durability of response (PFS) is inferior, likely because adverse events hinder the sustained administration of combination therapy. Thus, we do not currently believe that different ways of measuring response explain the difference between animal models and human trial data.

The most obvious difference between pre-clinical analysis of ICIs and clinical trials is that the former are performed primarily in syngeneic and genetically engineered mouse models, each of which is homogenous genetically (or nearly so). In contrast, patient populations are genetically heterogeneous and exhibit high variability in drug response. As a result, even if drug interactions identified in pre-clinical studies do occur in humans, too few patients are likely to receive sufficient benefit from both drugs for additivity or synergy to be manifest. A corollary of this hypothesis is that pre-clinical testing of ICI combinations would benefit from analysis in heterogeneous panels of tumor models. This has become routine in the case of patient-derived xenograft (PDX) mouse models and targeted anti-cancer therapies.^44^ However, animals used for traditional PDX modeling are immunocompromised and cannot be used to evaluate ICIs; generating panels of genetically heterogeneous mouse models with functional immune systems remains a daunting challenge.

What are the current and future implications of our findings? First, calculating the benefits from drug independence provides a sound strategy for designing new ICI combinations with predictable activity, which is desirable as the number of possible combinations grows. Conversely, if the predicted benefits of independence are insufficient for a new combination to exceed standard-of-care, the combination is likely to fail; this observation could help drug developers with portfolio prioritization. As monotherapy data become available for stratified subgroups (e.g. PD-L1 status, prior treatment, and patient demographics) it may be possible to predict combination therapy activity for specific patient subsets. Adverse effects remain the primary unknown, and better methods for predicting, preventing or mitigating toxicity are therefore a priority. Second, concurrent administration of independent drugs is not strictly necessary for benefit under the assumption of independence, and evaluation of sequential treatment or biomarker-guided choice of treatments is potentially warranted. However, in patients with rapidly progressing disease for whom second-line therapy may not be an option (due to declining functional status), using drugs in combination provides more chances for effective treatment upfront.

How can we move beyond independent drug action and realize the benefits of drug synergy? We hypothesize that the key is better patient stratification. If we can identify patients who respond to all or the majority of drugs in a combination^45^, we are likely enrich for drug additivity or synergy. It may be possible to develop predictive biomarkers for each therapy based on pre-treatment state, similar to those for ICI monotherapies^46,47^. An alternate approach that requires less knowledge about mechanism is to start patients on a multi-drug combination and then use pharmacodynamic biomarkers and on-treatment biopsies (or even pharmacokinetic data) to guide withdrawal of one or more inactive drugs and continuation or dose escalation of active ones. In either case, the discovery that existing ICI combinations operate via independent action serves to emphasize the importance of greater precision in cancer therapy based on better response biomarkers.

## METHODS

### Data sources

We sought data for all combination therapies with ICIs that were Food and Drug Administration (FDA)-approved as of April 2020. To assess whether a combination of therapies produces ‘additive or synergistic’ efficacy, we required Kaplan-Meier plots of PFS for the combination therapy, and for the different therapies comprising the combination, in matching patient cohorts. Several trials tested combinations of three or more drugs, where a standard-of-care combination (therapies ‘A+B’) was compared with a combination of more therapies (‘A+B+C’). In such cases the control arm combination (A+B) is regarded as one treatment; our analysis therefore assessed interactions between that standard treatment and the new therapy (‘C’), and is agnostic to drug interactions within control arms. Thus ‘single-agent response’ in some cases refers to a response to a previously established combination (this distinction is made clear in **Figure 2**).

Some trials randomized patients to three treatment arms, spanning two monotherapy treatments and their combination, and therefore contain all data needed for analysis. Other trials were lacking at least one monotherapy arm. In such cases we searched publications and conference proceedings for a clinical trial of that therapy meeting the following criteria: (i) patients had the same type and stage of disease (ii) patients were treated at the same or near-identical dose, and (iii) treatments involve the same line of therapy (for all combinations analyzed in this paper this was first-line). In 5 of 26 monotherapy arms, data was only available in previously-treated patients (second or third line). The impact of this discrepancy is addressed in the discussion. Complete treatment details and patient characteristics from the trials analyzed are presented in **Supplementary Table 1** and **Supplementary Table 2**.

Ultimately we found suitable combination and control monotherapy data for all but 3 of the 14 ICI combinations approved by the FDA (**Supplementary Table 3**). Two of the omitted combinations are nivolumab plus ipilimumab for renal cell cancer^48^, and nivolumab plus ipilimumab for microsatellite unstable or mismatch repair deficient metastatic colorectal cancer^49^; in both cases monotherapy data is unavailable making conclusions about drug interaction impossible. The third exception is lenvatinib plus pembrolizumab for advanced endometrial cancer^50^; we did not include this trial in the main text due to a shortage of pembrolizumab monotherapy data, and mismatching monotherapy cohorts, but a limited analysis is provided in **Supplementary Figure 2**.

We also included three ICI combination therapies lacking FDA approval where complete combination and monotherapy data was available for analysis: (i) pembrolizumab plus chemotherapy for advanced gastric and gastroesophageal junction cancers^18^ (ii) ipilimumab plus dacarbazine for metastatic melanoma^20^, and (iii) pembrolizumab plus dabrafenib and trametinib for metastatic BRAF-mutant melanoma^21^. The latter case was a notable exception from our requirement for near-identical dosing between arms, as most patients receiving ‘triple combination therapy’ for BRAF-mutant melanoma required dose reductions or interruptions to ameliorate treatment-related adverse events, the effect of which is discussed in the article.

### Data extraction

Published Kaplan-Meier curves of Progression Free Survival were opened in Adobe Illustrator to remove extraneous labels, and to produce separate figures for each treatment arm. Figures that were only available in rasterized form were digitally traced in Adobe Photoshop. Kaplan-Meier curves were digitized in Wolfram Mathematica 12 using published algorithms^6^. For Hazard Ratio calculations, individual patient data were reconstructed from Kaplan-Meier functions and ‘At Risk’ tables using published methods^26^.

### Simulation of independent action

As previously described^6^, Progression Free Survival (PFS) expected for the null hypothesis of independence was computed by randomly sampling data pairs of (PFStherapyA, PFStherapyB) from a joint distribution constructed from the clinically observed single-agent distributions, and assigning each simulated patient the better of their two PFS values (maximum of PFStherapyA, PFStherapyB). These joint PFS distributions were constructed with correlations of ρ=0.3±0.2, which accounts for partial cross-resistance between therapies. As reported^6^, this degree of cross-resistance was informed by drug responses in a large cohort of tumor xenografts, and is broadly consistent with clinical observations from sequential monotherapy trials and from comparing response rates at first and second line^5,28,31–33,51^. Partial correlations in drug response could in principle arise both from tumor-intrinsic properties and from patient status (e.g. age and co-morbidities). Simulations are performed with ≥10^4^ simulated patients in order to overcome sampling noise and produce consistent distributions, whereas stochastic variation is evident when simulating smaller cohorts. Identical predictions, which are fully deterministic, can be calculated from the equation

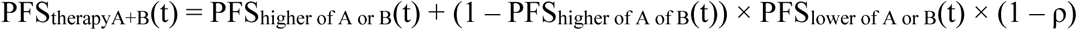

where *t* is time, and the calculation must be performed at each time point (e.g. in 0.05 month increments) to compute the full Kaplan-Meier curve. This is effectively Frei’s 1961 calculation^5^ with an adjustment for cross-resistance (ρ) and applied to time-series survival data. Additivity or synergy is supported when the efficacy of a combination therapy violates (by surpassing) the null hypothesis of independent action. Supplementary Code provides software and source data to execute these simulations for all trials analyzed here.

### Applicability of independent action model to PFS in advanced cancers, not adjuvant therapy, indolent tumors, or Overall Survival data

The calculation of independent action applies to situations for which an observed rate of tumor control (Progression Free Survival) can be attributed to the effect of treatment. This calculation would not be valid in scenarios where the ‘survival’ percentage does not reflect the percentage of patients in which treatment has prevented tumor growth up to the time point in question. Thus, valid modeling scenarios include PFS for advanced cancers that will grow (progress) if they are not treated, for time points after a patients’ first or second scan (before that, there is likely to have been little opportunity to observe progression). This is demonstrated by analysis of clinical trials in advanced cancers that included a ‘placebo-only’ arm, in which large majorities of patients exhibit progression before 4 months (**Supplementary Figure 4**). Logically, combining two placebos should provide no benefit. To apply our modeling approach to this scenario recall that any treatment is ‘cross-resistant’ with itself, resulting in ρ=1, in which case independent action expects no benefit from combining a treatment with itself. Because ‘placebo + placebo’ combination is the same ‘treatment’ given twice, it provides no benefit in the independent action model implemented in this paper. Alternatively, it is possible to take an empirical approach and treat placebos like any other therapy described in this paper and being subject to independent modeling (with ρ=0.3±0.2). Under this assumption, the benefit predicted for placebo plus placebo arms at 12 months ranges between 0% and 1.5% for five trials involving different types of cancer (average 0.6%; **Supplementary Figure 4**). This is negligible when compared to the degree of benefit observed in successful ICI trials.

Several clinical scenarios cannot or should not be analyzed using the independent action model described here. These include ‘Disease Free Survival’ for adjuvant therapy following surgery with curative intent, because when some patients are cured by surgery prior to therapy, the DFS rate cannot be attributed to the efficacy of adjuvant therapy (e.g. if surgery cures 50% of patients, and surgery plus adjuvant therapy cures 60%, one cannot conclude that adjuvant therapy is effective in 60% of patients). Indolent or benign tumors, often subject to a ‘watch and wait’ approach, are also invalid scenarios because PFS may be high and long-lasting with no therapy. Finally, the independent action calculation should not be applied to Overall Survival (OS) data, because OS can remain high even after ineffective therapy, and post-progression survival has many influences aside from the therapy assigned in a trial, including second-line or salvage therapies that may be used following progression. Empirically, in trials that include a ‘placebo only’ arm, OS distributions can show a substantial tail (**Supplementary Figure 4**). Because OS percentages are systematically higher the percentage of patients in which therapy was effective (up to any given query time), the independent activity of a drug cannot be inferred from OS data, so the independent action simulation should not be applied to OS data.

## Data Availability

All source data is from published clinical trials cited in references and Supplementary Tables 1-3.

